# Partial Pressure of Oxygen-Guided Adrenal Venous Sampling in Primary Aldosteronism

**DOI:** 10.1101/2025.06.29.25330380

**Authors:** Kei Omata, Yuta Tezuka, Hiromitsu Tannai, Yoshikiyo Ono, Hiroki Kamada, Sota Oguro, Yoshihide Kawasaki, Akihiro Ito, Yuto Yamazaki, Hironobu Sasano, Takashi Suzuki, Tetsuhiro Tanaka, Kei Takase, Hideki Katagiri, Fumitoshi Satoh

**Affiliations:** Department of Diabetes, Metabolism, and Endocrinology, Tohoku University Graduate School of Medicine; Department of Diagnostic Radiology, Tohoku University Graduate School of Medicine; Department of Urology, Tohoku University Graduate School of Medicine; Department of Pathology, Tohoku University Graduate School of Medicine; Department of Nephrology and Hypertension, Tohoku University Graduate School of Medicine

## Abstract

**[Background]:** Primary aldosteronism (PA) is a leading cause of endocrine hypertension, characterized by the increased cardiovascular risks. Adrenal venous sampling (AVS) plays a pivotal role in optimization of treatment strategy for PA. However, technical challenges, such as anatomical variations and the small sizes of target vessels, often preclude us from appropriate cannulation to the adrenal veins. This study aimed to explore the distribution of partial pressure of oxygen (pO_2_) in the adrenal and their neighboring veins, inspired by our awareness of lighter red color of adrenal venous blood than the others.

**[Methods]:** We enrolled 168 PA patients who underwent AVS at our center from 2021 to 2024. During AVS, we collected residual blood samples (0.2 mL for each) from bilateral adrenal, hepatic, inferior phrenic and external iliac veins both at baseline and under cosyntropin stimulation to perform blood gas analysis. Statistical analysis was conducted to evaluate the pO_2_ distributions and its associations with clinical parameters.

**[Results]:** The pO_2_ levels were significantly higher in bilateral adrenal veins than in their counterparts. The pO_2_ levels tended to remain the same even after cosyntropin initiation, with its decrement only detected in the adrenal veins. In addition, the levels of partial pressure of carbon dioxide were lower in the adrenal veins than in the others. Of note, in the adrenal veins, pO_2_ levels were significantly negatively correlated with ipsilateral levels of both aldosterone and cortisol. Finally, results of our present study firstly demonstrated the significance of evaluating pO_2_ in identifying adrenal veins compatible to the selectivity index of cortisol, a standard criterion for successful cannulation.

**[Conclusion]:** This is the first study to examine the pO_2_ dynamics in the human adrenal and non-adrenal veins, demonstrating its potential as a reliable tool for improving AVS cannulation success rates. Our findings also presented the oxygen consumption in the adrenal glands for steroidogenesis during cosyntropin stimulation. The pO_2_ measurement is a faster, easier and less-expensive tool enhancing AVS techniques. Our findings could improve clinical practice and research towards the next stage of adrenocortical endocrinology.

**Clinical Perspective:** *What is new?:* -The adrenal venous specimens appear lighter red than those from non-adrenal veins, which is considered to be due to significantly higher partial pressure of oxygen in the adrenal veins. -Cosyntropin stimulation results in decreased partial pressure of oxygen in the adrenal veins, indicating increased adrenal consumption of oxygen due to ACTH-driven steroid synthesis.

*What are the clinical implications?:* -Measurement of partial pressure of oxygen has a high ability for identifying adrenal veins compatible to the selectivity index of cortisol, a standard criterion for successful adrenal vein cannulation. -Venous blood gas analysis is a faster, easier and less-expensive tool for confirmation of successful cannulation to the adrenal veins when performing adrenal venous sampling.

## Introduction

Primary aldosteronism (PA) is a leading cause of endocrine hypertension, accounting for 3-12% of all patients with hypertension in primary care settings^1^. A pile of evidence elucidated that PA frequently triggered cardiovascular disease (CVD) in both blood pressure (BP)- and mineralocorticoid-dependent manners: the enhanced risks of CVD in PA were reported as 1.67-1.77, 2.05, 2.03-2.58 and 2.68 times higher for coronary artery disease, heart failure, stroke and proteinuria, respectively, compared to those with essential hypertension (EH)^2–4^. In addition, PA could lead to a higher risk of mortality than EH^5^. PA-specific treatment, therefore, plays a pivotal role in mitigating the increased CVD risks as well as achieving better BP control^6^. Results of one meta-analysis revealed that adrenalectomy for unilateral PA resulted in 39% and 99% of complete clinical and biochemical outcomes, respectively, based on the PASO criteria^7^. Administration of mineralocorticoid receptor antagonists (MRAs) could also reverse the excess risks of developing comorbidities in bilateral PA, while the efficacy of medical treatment depends on MRA titration by physicians. Several clinical studies also reported medically-treated PA patients harboring persistent renin suppression had higher CVD incidences than those who achieved normal or elevated renin status after therapy^8, 9^. Therefore, surgically-treated PA patients tend to have reduced events of major adverse CVD (odds ratio [OR], 0.55), atrial fibrillation (OR, 0.35) and mortality (OR, 0.33) than medically-treated patients^10, 11^. Therefore, whether treatment is optimized for each PA subtype or not has a great impact on healthy life expectancy in the patients.

Toward classification of PA patients, adrenal venous sampling (AVS) has been employed as a gold standard procedure to differentiate unilateral PA, surgery-curable form, from bilateral PA^6^. The techniques of AVS have been well developed, but still challenging due to the difficulties in cannulating adrenal veins. In fact, the success rate of bilateral adrenal venous cannulation was reported as 60-80% of all AVS cases^12–14^. A major cause of AVS failure is the difficulty in intraprocedural confirmation of adrenal venous cannulation. A couple of studies reported anatomical variation of right and left adrenal veins (RAV and LAV)^15, 16^. The RAV normally flows into the inferior vena cava (IVC), whereas we reported that 20% of PA patients who underwent AVS had a common trunk of RAV and the accessory hepatic vein (HV)^15^. Those cases demonstrated various locations of the RAV orifice and angles between RAV and HV or IVC. The close proximity of the openings of the RAV and HV could therefore prevent successful RAV cannulation. Furthermore, on venograms, both veins could exhibit similar morphologies, leading to misidentification and sampling from the HV instead of the RAV. This is considered one of the causes of AVS failure. On the other hand, the LAV has been reported to form a common trunk with the inferior phrenic vein (IPV), draining blood from the diaphragm and stomach, before LAV flows into the left renal vein in more than 90% of the relevant cases^16^. Those anatomical features and individual differences sometimes resulted in misidentification of adrenal veins even in the experienced centers. Today, the selectivity index using cortisol levels in the target and the reference veins is globally used to judge whether adrenal veins are appropriately cannulated or not^17^. In order to improve the success rate of AVS, intraprocedural measurement of cortisol has been suggested in addition to preoperative imaging focusing on adrenal veins^18, 19^.

Recently, we have noticed the macroscopic color difference between blood specimens from the external iliac vein (EIV) and the adrenal vein in AVS. The red color of adrenal venous blood appeared to be lighter than those of the neighboring veins, including HV and EIV. Subsequent evaluation revealed that partial pressure of oxygen (pO_2_) was higher in adrenal venous samples than in other venous samples, including HV and IVC. We, therefore, conducted the present study to prospectively investigate the pO_2_ differences among blood obtained from adrenal veins and nearby veins, with a hypothesis that the pO_2_ measurement is an indicator for successful cannulation of adrenal veins during the AVS procedure.

## Methods

### Ethical considerations

This study protocol was developed in accordance with the tenets of Declaration of Helsinki and approved by Tohoku University Hospital institutional review board (2023-1-817). We obtained written informed consents from all the participants for the procedures for this study.

### Eligibility of the participants for this study

In order to evaluate pO_2_ differences among adrenal and the relevant veins, we included PA patients who underwent AVS for subtyping in our center from April 2021 to March 2024. Diagnosis of PA was performed as per the PA guidelines^20, 21^. Briefly, hypertensive patients underwent evaluation of plasma renin and aldosterone after sufficient withdrawal of antihypertensive agents which interfere renin-angiotensin-system. Blood samples for renin and aldosterone evaluation were obtained after 30 minutes rest at supine position in the morning. Screening criteria for PA were defined as a higher plasma aldosterone concentration (PAC) than 6 ng/dL along with renin suppression (plasma renin activity [PRA] <1 ng/mL/h or its concentration [PRC] <4 pg/mL)^22^. Both PAC and PRC were measured by the recently developed chemiluminescent enzyme immunoassay^22^, while PRA by commercially available enzyme immunoassay. The diagnosis of PA was established based on the results of captopril challenge test (aldosterone-to-renin ratio >8.2 ng/dL per ng/mL/h after 50 mg captopril loading) and/or saline infusion test (PAC >1.2 ng/dL after 2 L saline injection)^20, 23^. Those who had overt subclinical Cushing syndrome or other uncontrolled endocrinopathies, including thyroid dysfunction, were excluded for this study^24^. Blood pressure was measured after 5 minutes of rest with HEM 7120 (Omron Heathcare Co Ltd, Kyoto, Japan). Basic clinical information of the participants were collected by reviewing their medical records.

### Adrenal Imaging by computed tomography

Non-contrast and four-phase dynamic contrast-enhanced computed tomography (CT) was primarily employed to evaluate adrenal nodules, adrenal veins and their neighboring veins using a 160-row multidetector CT system (Aquilion Precision; Canon Medical Systems, Otawara, Japan)^15^. The tube voltage was set to 120LkVp and the tube current to 310LmA. The scanner operated with a rotation time of 0.5 seconds and a pitch of 0.8. Transverse reconstructed images were acquired with a slice thickness and slice interval of 1 mm, a matrix size of 512L×L512 pixels, and a field of view of approximately 320 mm for all phases. Specifically, for the late arterial phase images, higher resolution was achieved with a slice thickness and interval of 0.25 mm, a matrix size of 512L×L512 pixels, and a field of view of approximately 240 mm. A non-ionic contrast agent, containing 300–370Lmg iodine/mL, was injected at a dose of 600 mg I per patient body weight into a peripheral vein over 25 seconds. When the attenuation value reached a preset threshold (CT value on plain CT plus 50 Hounsfield Units) at the level of the abdominal aorta, early arterial-phase scanning automatically commenced^15^. Late arterial-phase scanning began 13 seconds after the completion of the first scan. Venous-phase and delayed-phase scanning started 70 seconds and 3 minutes, respectively, after the initiation of contrast injection.

### Subtyping using segmental AVS

The procedure was performed by interventional radiologists, with endocrinologists in attendance. A 5LF or 7LF sheath was inserted into the bilateral femoral veins under ultrasonography guidance. A 5LF or 6.5LF diagnostic catheter for the left or right adrenal veins (Adselect, Hanaco Medical, Tokyo, Japan; MK adrenal type; Hanaco Medical, Tokyo, Japan) and a 2–2.9LF split-tip microcatheter (Goldcrest Neo microcatheter type OM, Medico’s Hirata, Osaka, Japan) were also utilized^25–27^. First, the LAV was cannulated with the catheter. Subsequently, the HV and RAV were cannulated with right adrenal catheter. Blood sampling was carefully conducted to obtain a minimum of 1.5 mL from RAV, HV and LAV (specially, from sites before and after IPV confluence, referred to LAV_CV_ and LAV_IPV_, respectively.) and from right EIV (reference) using a microcatheter as needed. HV sampling was primarily performed from the middle or right HV, and occasionally from an accessory HV. Venous samples were also collected from the IPV when adrenal blood flow to the IPV was suspected. Bilateral AVS was completed within a short time frame. Subsequently, a 200Lμg intravenous bolus of cosyntropin (synthetic adrenocorticotropic hormone, ACTH) was administered, followed by a continuous infusion of 50 μg/h via a peripheral vein 30 minutes later^25–28^. Fifteen minutes after the initiation of ACTH stimulation, sampling from the same sites and bilateral adrenal tributary veins was conducted. Heparinization was performed to prevent coagulation inside the catheter. Basically, oxygen was administrated at 2 L/min throughout the AVS procedure to keep oxygen saturation over 98%.

The laterality of PA was determined based on the lateralized index under ACTH stimulation as previously reported^28^. Briefly, unilateral PA was identified in the case where the lateralized index, a quotient of the aldosterone/cortisol ratio obtained from the adrenal central vein of the dominant side divided by that of the other side, was higher than 2.6. Some cases were also categorized as unilateral PA by segmental AVS when the aldosterone/cortisol ratios were similar in bilateral adrenal central veins but significantly elevated only in a specific segment, often corresponding to a CT-detectable tumor. Successful cannulation to the adrenal veins were confirmed based on the selectivity index (the ratio of the cortisol level in the target vein divided by that of EIV >5) as well as angiography during AVS.

### Measurement of steroids and partial pressure of oxygen and carbon dioxide in AVS samples

As routine diagnostic procedures of AVS, PAC and serum cortisol were measured employing commercially available assays, the chemiluminescent enzyme immunoassay (Lumipulse Presto Aldosterone; Fujirebio Inc., Tokyo, Japan) and the electro chemiluminescent immunoassay (Elecsys Cortisol-II; Roche Diagnostics, Rotkreuz, Switzerland), respectively. Assessment of both hormones was performed using blood samples from the adrenal veins and EIV, while only cortisol levels were measured in HV and IPV samples. For this study, we also performed blood gas analysis in residual venous samples collected from both adrenal veins, HV, IPV and EIV during AVS. The pO_2_ and partial pressure of carbon dioxide (pCO_2_) were measured employing ABL800 FLEX (Radiometer Medical A/S, Bronshoj, Denmark). Blood gas analysis was performed with 0.2 mL of residual samples immediately after collection of the blood samples during AVS. Gas analysis within 3 minutes confirmed that pO_2_ stability remained within a 2% variation (**Figure S1**).

### Statistical analysis

We employed SPSS (version 28.0, Armonk, NY: IBM Corp) for all statistical analyses performed in this study. For clinical information, parametric and non-parametric variables were shown as mean ± standard deviation and median [an interquartile range], respectively. The comparison of blood pO_2_ or pCO_2_ levels between two veins or before and after ACTH stimulation was performed with the Wilcoxon signed-rank test. In cases with missing values, we excluded the cases with missing data only for the specific statistical analysis affected. The differences of pO_2_ or pCO_2_ between a target vein and EIV were shown as ΔpO_2_ or pCO_2_, respectively, by subtracting their levels at EIV from those at a target vein. In addition, we employed Spearman’s correlation to reveal the associations between venous gas status and steroid production. Subsequently, the influence of clinical parameters on pO_2_ levels was assessed with stepwise regression. Finally, receiver operating characteristic (ROC) curves were constructed to investigate the discriminative ability of pO_2_ and pCO_2_ levels in identifying adrenal veins, based on the cases in which target parameters were available for both the adrenal veins and their counterpart veins. The optimal cut-off value for ROC analysis was determined based on the Youden index. Statistical significance was set at p < 0.05 for all the statistical analysis performed in this study.

## Results

We enrolled 179 PA patients whose baseline characteristics were summarized in **Table S1**. Overall, 89 (49.7%) were men and the mean age and body mass index were 53.0 years old and 24.9 kg/m^2^, respectively. Adrenal imaging found unilateral and bilateral adrenal tumors in 100 (55.9%) and 17 (9.5%) patients, respectively. A combination of adrenal imaging and AVS confirmed the confluences of HV and RAV, and IPV and LAV in 30 (16.8%) and 176 (98.3%) of the patients, respectively. Based on segmental AVS, 104 (58.1%) patients were diagnosed with unilateral PA. Of these, 81 (77.9%) showed concordant laterality of hyperaldosteronism between adrenal imaging and segmental AVS, while others had bilateral adrenal tumors or no tumor on adrenals. Of the 179 PA patients, 11 (6.1%) didn’t receive oxygen support during AVS. The subsequent analysis focusing on pO_2_ status was, therefore, limited to 168 patients who underwent AVS with oxygen supplementation to ensure accurate evaluation.

### Blood gas evaluation of venous samples obtained during AVS

#### 1. Venous pO_2_ levels before ACTH stimulation

A representative image illustrating the color difference between blood specimens from the EIV and the adrenal vein is shown in **Figure 1**. First, we measured blood pO_2_ in the adrenal and neighboring veins without ACTH stimulation (**Figure 2A and Table S2**). Among 168 PA patients, baseline blood samples from RAV, HV, EIV, IPV, LAV_IPV_ and LAV_CV_ were available for gas analysis in 158 (94.0%), 165 (98.2%), 168 (100%), 73 (43.4%), 86 (51.2%) and 166 (98.8%) cases, respectively. Consistent with the lighter red blood from the adrenal veins, both blood pO_2_ levels were significantly higher in RAV and LAV_CV_ than in EIV (66.3 [59.3, 76.1] and 66.5 [57.5, 76.2] mmHg vs. 43.2 [40.0, 47.0] mmHg, respectively; *p* <0.0001 for both). There was no significant difference of blood pO_2_ between RAV and LAV_CV_ (*p*=0.07). Among the limited cases with available gas analysis in the adrenal tributary vein, there was no significant difference of pO_2_ levels between the adrenal central and tributary veins (**Table S3**). Similarly, pO_2_ levels in RAV and LAV_CV_ were significantly higher than those in their neighboring veins, HV and IPV, respectively (40.8 [37.6, 44.3] and 43.1 [35.5, 51.7] mmHg in HV and IPV, respectively; *p* <0.0001 for both) (**Table S2**). Moreover, pO_2_ levels were lower in HV than in EIV (*p* <0.0001), while there was no significant difference of pO_2_ between IPV and EIV. Thus, blood pO_2_ levels in LAV decreased after IPV confluence (66.5 [57.5, 76.2] mmHg in LAV_CV_ vs. 56.0 [50.3, 62.5] mmHg in LAV_IPV_, *p* <0.0001). The differences of pO_2_ levels between the target veins and EIV,ΔpO_2_, were summarized in **Figure 2B**. The ΔpO_2_ clearly demonstrated that blood pO_2_ levels in RAV were higher than those in EIV in all available cases (**Figure 2B**). In addition, 164 out of 166 (98.8%) cases showed similar results between LAV_CV_ and EIV pO_2_ levels.

**Figure 1.**
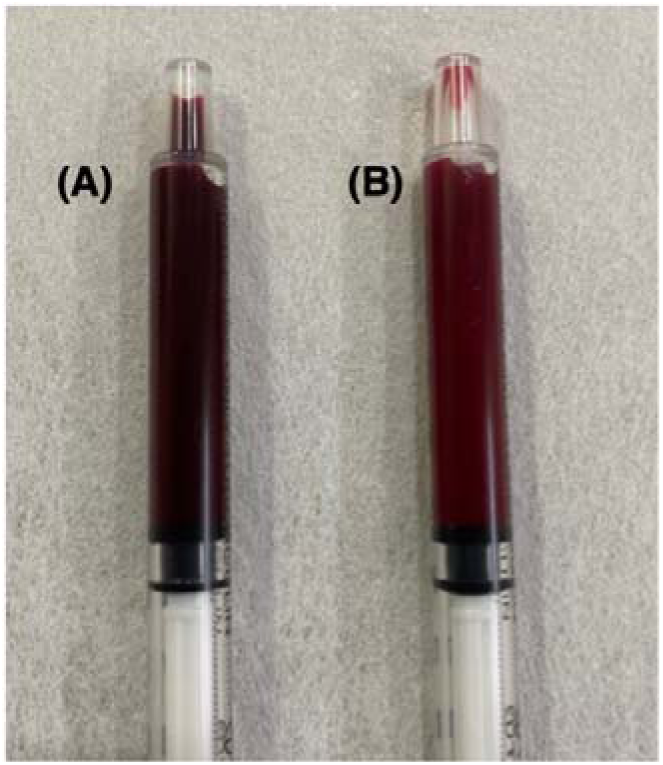
Different blood colors of the external iliac vein (A) and the adrenal vein (B) obtained during adrenal venous sampling. Adrenal blood showed brighter red than the others.

**Figure 2.**
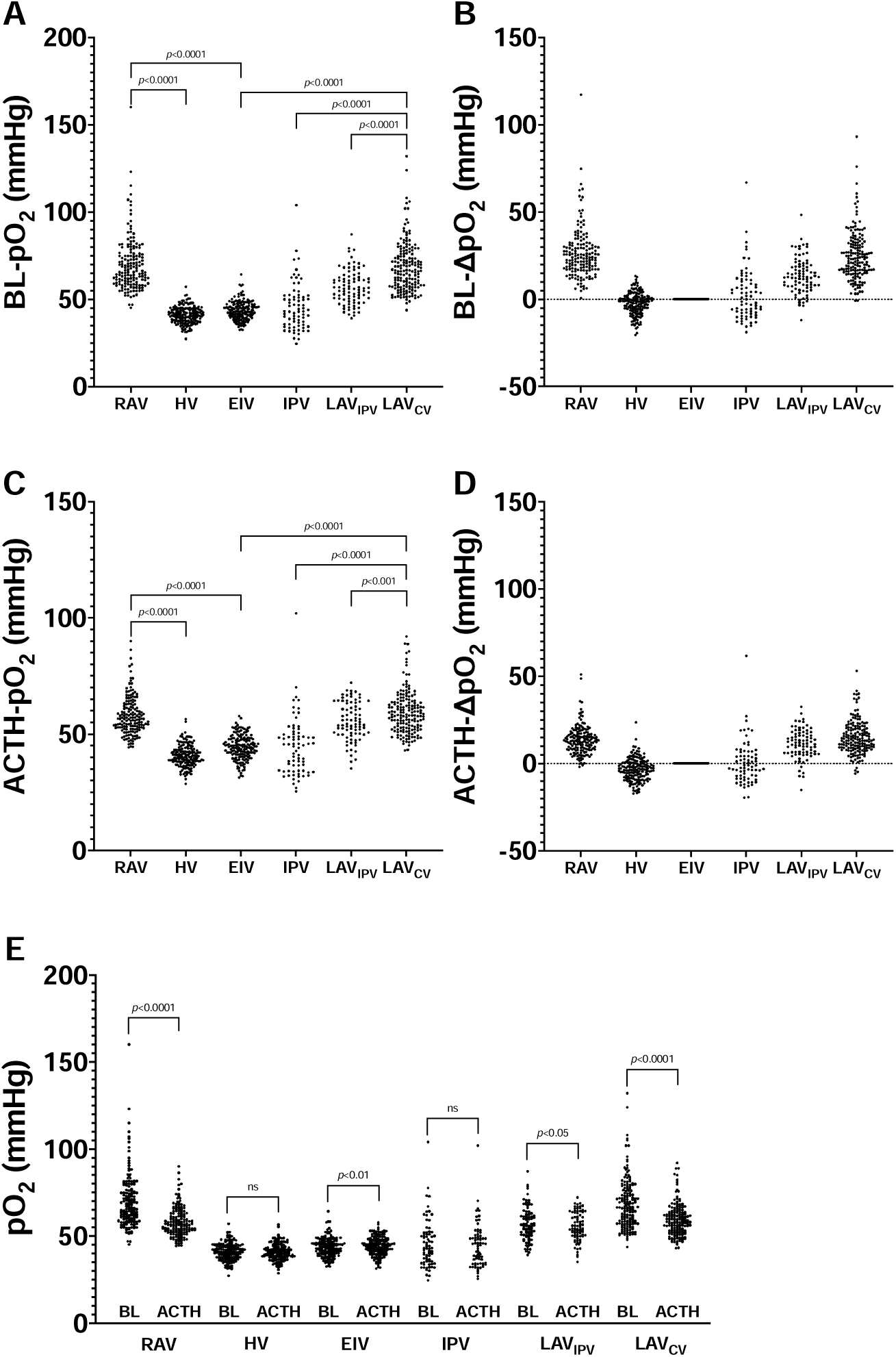
The pO_2_ evaluation in the adrenal veins and their surrounding veins. pO2, partial pressure of oxygen; RAV, right adrenal vein; HV, hepatic vein; EIV, external iliac vein; IPV, inferior phrenic vein; LAV_IPV_, left adrenal vein after the confluence of IPV; LAV_CV_, the central vein of left adrenal; BL, baseline; ACTH, cosyntropin stimulation. Δ pO2 was calculated by subtracting pO_2_ levels at EIV from those at a target vein. Baseline pO_2_ levels and calculated Δ pO_2_ in the examined veins are shown in panel A and B. The pO_2_ levels under cosyntropin stimulation and Δ pO_2_ are depicted in panel C and D. The pO_2_ levels are compared between baseline and under cosyntropin stimulation in panel E. ns, not significant.

#### 2. Changes of venous pO_2_ levels after ACTH stimulation

Then, we evaluated blood pO_2_ levels in the same veins under ACTH stimulation in 167 available cases (**Figure 2C and D, and Table S2**). Blood pO_2_ levels significantly decreased in RAV, from 66.3 [59.3, 76.1] mmHg to 56.6 [52.7, 62.8] mmHg (*p* <0.0001), and LAV_CV_, from 66.5 [57.5, 76.2] mmHg to 58.2 [52.7, 63.8] mmHg (*p* <0.0001) after ACTH injection (**Figure 2E**). In contrast, the pO_2_ levels didn’t change in either HV, from 40.8 [37.6, 44.3] mmHg to 40.4 [37.8, 43.4] mmHg (not significant), or IPV, from 43.1 [35.5, 51.7] mmHg to 45.1 [34.3, 50.0] mmHg (not significant: **Figure 2E**). In EIV, blood pO_2_ levels slightly increased from 43.2 [40.0, 47.0] mmHg to 44.4 [41.2, 47.8] mmHg (*p* =0.007). However, the dominance of blood pO_2_ levels in the adrenal veins over those in examined non-adrenal veins was maintained even under ACTH stimulation (**Figure 2D**).

Those trends of pO_2_ distribution at baseline and after ACTH stimulation were similar even in 11 PA cases where O_2_ was not administered during AVS (**Figure S2**). However, there were significant differences of pO_2_ levels between cases with and without oxygen administration, particularly in those of adrenal veins at baseline (**Table S4)**.

#### 3. Evaluation of venous pCO_2_ levels before and after ACTH stimulation

Notably, blood pCO_2_ levels were significantly lower in the adrenal veins compared to their neighboring veins (**Figure 3A and B**). At baseline, the pCO_2_ levels in RAV were lower than those in EIV and HV, measuring 41.6 [39.0, 43.7] mmHg in RAV versus 46.5 [43.7, 48.4] mmHg in EIV and 43.4 [40.8, 45.7] mmHg in HV (*p* <0.0001 for both). Similarly, the pCO_2_ levels were lower in LAV_CV_ than in EIV and IPV, with values of 42.2 [39.9, 44.3] mmHg in LAV_CV_ compared to 46.5 [43.7, 48.4] mmHg in EIV and 44.7 [42.6, 46.9] mmHg in IPV (*p* <0.0001 for both). Following ACTH stimulation, blood pCO_2_ levels significantly increased to 43.4 [41.3, 45.5] and 43.5 [41.2, 46.0] mmHg in RAV and LAV_CV_, respectively (*p* <0.0001 for both). In addition, the initiation of ACTH resulted in a mild elevation of blood pCO_2_ levels in HV, from 43.4 [40.8, 45.6] mmHg to 44.2 [41.1, 46.3] mmHg (*p* =0.02), and IPV, from 44.7 [42.5, 46.9] mmHg to 45.2 [42.9, 48.2] mmHg (*p* =0.001), but not EIV (**Figure 3B**).

**Figure 3.**
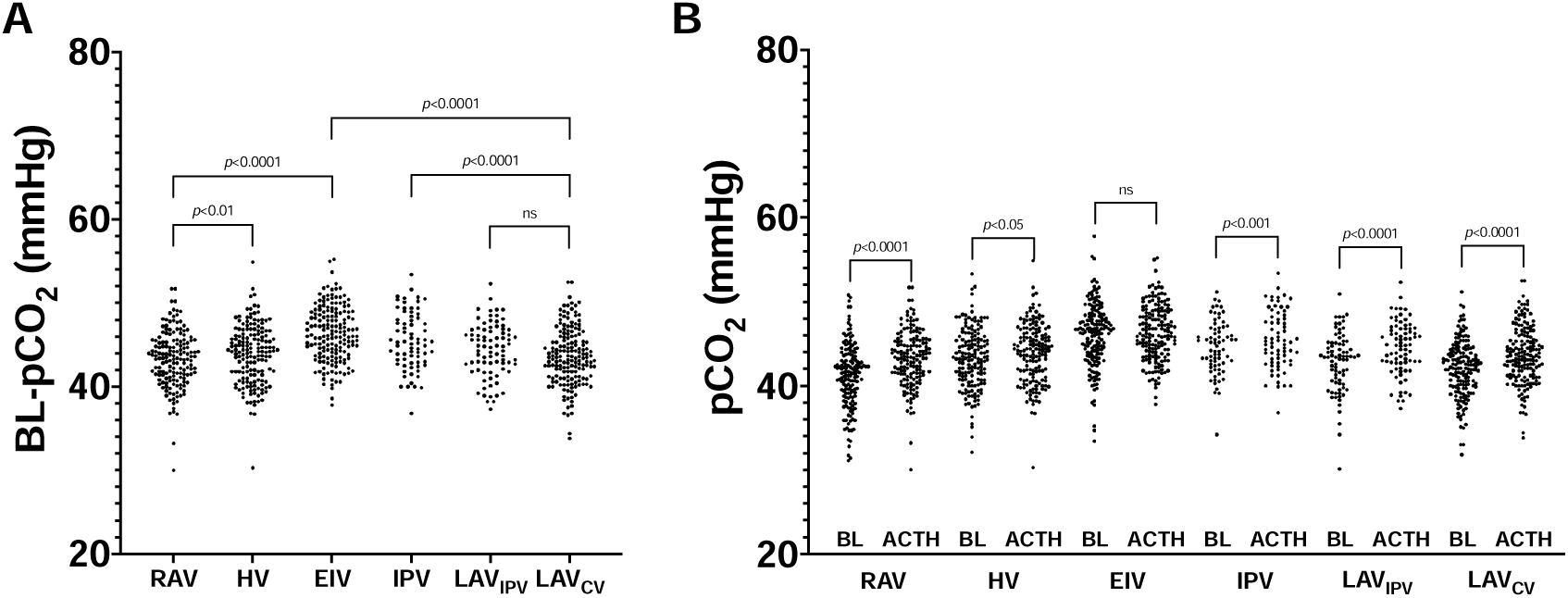
The pCO_2_ evaluation in the adrenal veins and their surrounding veins. pCO_2_, partial pressure of carbon dioxide; RAV, right adrenal vein; HV, hepatic vein; EIV, external iliac vein; IPV, inferior phrenic vein; LAV_IPV_, left adrenal vein after the confluence of IPV; LAV_CV_, the central vein of left adrenal; BL, baseline; ACTH, cosyntropin stimulation. Baseline pCO_2_ levels in each vein are shown in panel A. Changes of pCO_2_ levels after cosyntropin injection are summarized in panel B. #, *p*<0.05; **, *p*<0.01; ***, *p*<0.001; ****, *p*<0.0001; ns, not significant.

#### 4. Influential factors on venous gas status in adrenal veins

Intriguingly, in both adrenal veins, cortisol secretion was negatively correlated with pO_2_ levels, both at baseline and under ACTH administration (**Figure 4A-F**). At baseline, the correlation between cortisol secretion and pO_2_ was weak (Spearman r =-0.2265 and -0.2588, *p* =0.004 and 0.0008 in RAV and LAV_CV_, respectively) (**Figure 4A and 4D**), but became moderate after ACTH stimulation (Spearman r =-0.5142 and -0.5347, *p* <0.0001 for both in RAV and LAV_CV_, respectively) (**Figure 4B and 4E**). The cortisol increase, calculated as cortisol levels after ACTH stimulation divided by baseline levels, also showed a negative correlation with pO_2_ changes between before and after ACTH initiation (Spearman r =-0.2645 and -0.2529, *p* =0.0009 and 0.001 in RAV and LAV_CV_, respectively) (**Figure 4C and 4F**). Similarly, PAC was also negatively correlated with pO_2_ levels (**Figure 4G-L**). The correlations of pO_2_ with PAC were similar to those with cortisol levels at baseline (Spearman r =-0.2221 and -0.2975, *p* =0.005 and <0.0001 in RAV and LAV_CV_, respectively) (**Figure 4G and 4J**), but relatively weaker under ACTH stimulation (Spearman r =-0.2183 and -0.2891, *p* =0.006 and 0.0002 in RAV and LAV_CV_, respectively) (**Figure 4H and 4K**). The aldosterone increase, calculated as PAC after ACTH stimulation divided by baseline levels, was also weakly but negatively correlated with the pO_2_ changes (Spearman r =-0.1728 and -0.2087, *p* =0.03 and 0.007 in RAV and LAV_CV_, respectively) (**Figure 4I and 4L**). Furthermore, LAV_CV_ showed a negative correlation between aldosterone-to-cortisol ratios and pO_2_ (Spearman r =-0.2037 and -0.1525, *p* =0.009 and 0.05 at baseline and after ACTH stimulation, respectively), but not RAV (not significant). On the other hand, HV showed a positive correlation between cortisol levels and pO_2_ only at baseline (Spearman r =0.2101, *p* =0.007), and IPV showed no correlation between them (**Figure S3)**. In addition, no significant correlation was observed between pCO_2_ and other parameters above in both adrenal veins and non-adrenal veins.

**Figure 4.**
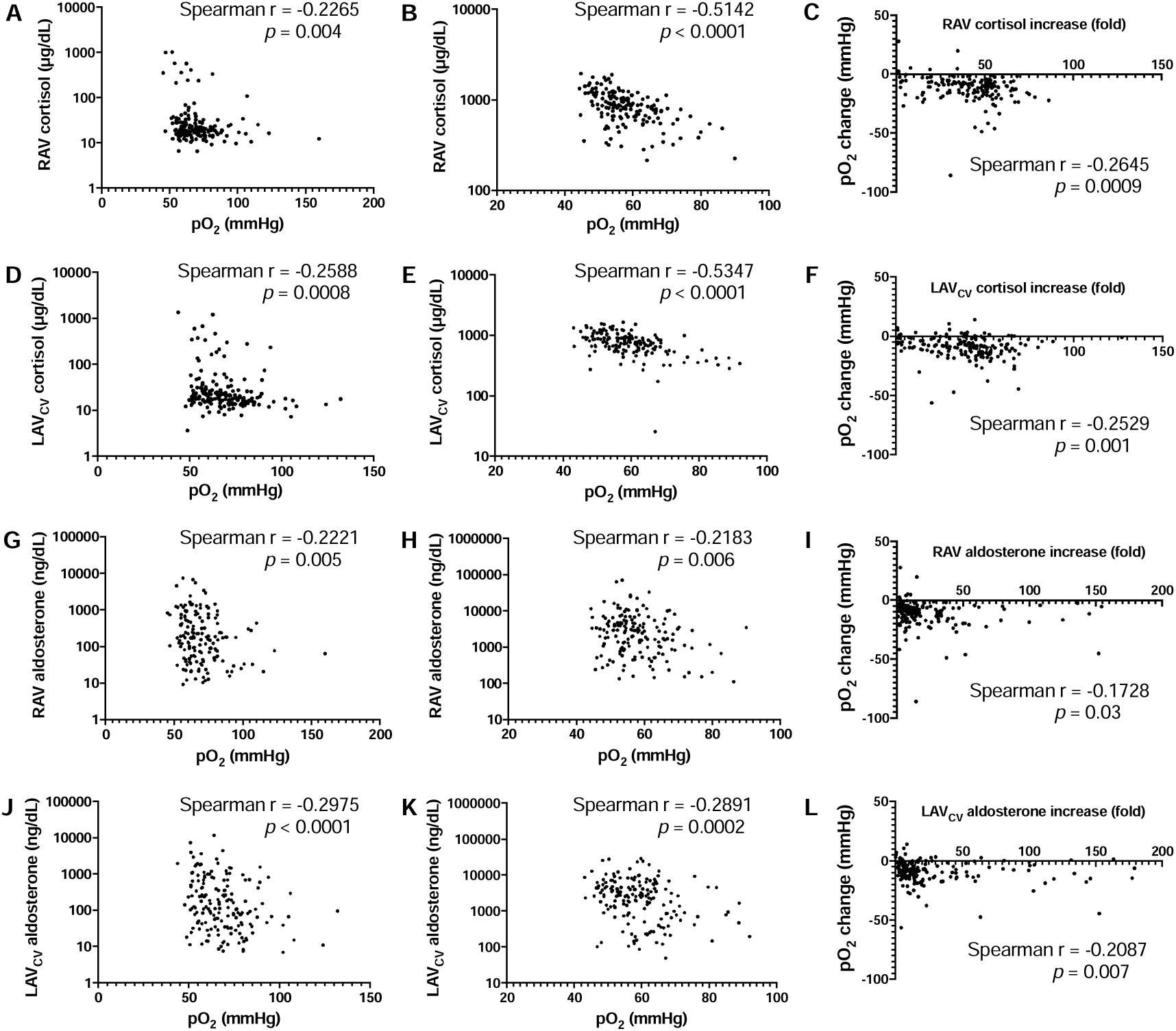
Correlations between pO_2_ and steroid production in the adrenal veins. pO_2_, partial pressure of oxygen; RAV, right adrenal vein; LAV_CV_, the central vein of left adrenal. The pO_2_ change was calculated by subtracting pO_2_ levels under cosyntropin stimulation from their baseline levels. Cortisol or aldosterone increase was represented by dividing cortisol or aldosterone levels following cosyntropin injection by their baseline levels. The correlations between pO_2_ and cortisol production in RAV and LAV_CV_ are shown in panel A-C, and D-F, respectively. Panel A and D show the baseline evaluation, while panel B and E illustrate the parameters under cosyntropin stimulation. Changes in pO_2_ and cortisol production after cosyntropin injection are displayed in panel C and F. For aldosterone, panel G-I and J-L represent the associations with pO_2_ for RAV and LAV_CV_, respectively. The associations between pO_2_ and aldosterone at baseline and following cosyntropin injection are shown in panel G and J, and H and K, respectively. Changes in pO_2_ and aldosterone production after cosyntropin injection are evaluated in panel I and L.

Finally, multiple regression analysis revealed that body mass index and ipsilateral PAC were negatively associated with pO_2_ levels in RAV and LAV_CV_, regardless of ACTH stimulation (**Table 5S**). Additionally, age and ipsilateral cortisol secretion were confirmed as influential factors on pO_2_ levels in LAV_CV_ after ACTH stimulation and RAV for both statuses. Neither PA laterality nor the presence of an adrenal tumor was associated with pO_2_ status.

### Application of blood gas analysis on detection of adrenal veins

#### 1. ROC analysis of venous pO_2_ in distinguishing adrenal veins

Finally, we investigated the discriminative ability of venous pO_2_ and pCO_2_ in identifying adrenal veins (**Figure 5**). At baseline, ROC curves demonstrated excellent performance of both actual pO_2_ and ΔpO_2_ in differentiating RAV from HV, comparable to the SI (**Figure 5A**). Optimal cutoff values for RAV detection based on the Youden index were determined as 1.64 (sensitivity 0.981, specificity 0.987), 50.9 mmHg (sensitivity 0.981, specificity 0.981) and 10.2 mmHg (sensitivity 0.942, specificity 0.981) for the SI, actual pO_2_ and ΔpO_2_, respectively (**Table 1**). For actual pO_2_, cutoff values of 45.2 and 57.3 mmHg showed a sensitivity of 1.000 and specificity of 1.000 for RAV detection, respectively. In addition, a higher ΔpO_2_ values greater than 0.6 and 13.5 mmHg were associated with a sensitivity of 1.000 and a specificity of 1.000 for RAV, respectively, as well. After ACTH stimulation, a higher SI exceeding 9.4 demonstrated a sensitivity and specificity of 1.000 for RAV detection (**Figure 5B**). The evaluation of actual pO_2_ and ΔpO_2_ also proved helpful for RAV cannulation even under ACTH stimulation (**Table 1**). An actual pO_2_ greater than 47.2 mmHg exhibited a sensitivity of 0.955 and specificity of 0.929, while a ΔpO_2_ greater than 3.9 mmHg demonstrated a sensitivity of 0.929 and specificity of 0.897. However, the performance of actual pO_2_ and ΔpO_2_ was inferior to that of the SI (*p*<0.01).

**Figure 5.**
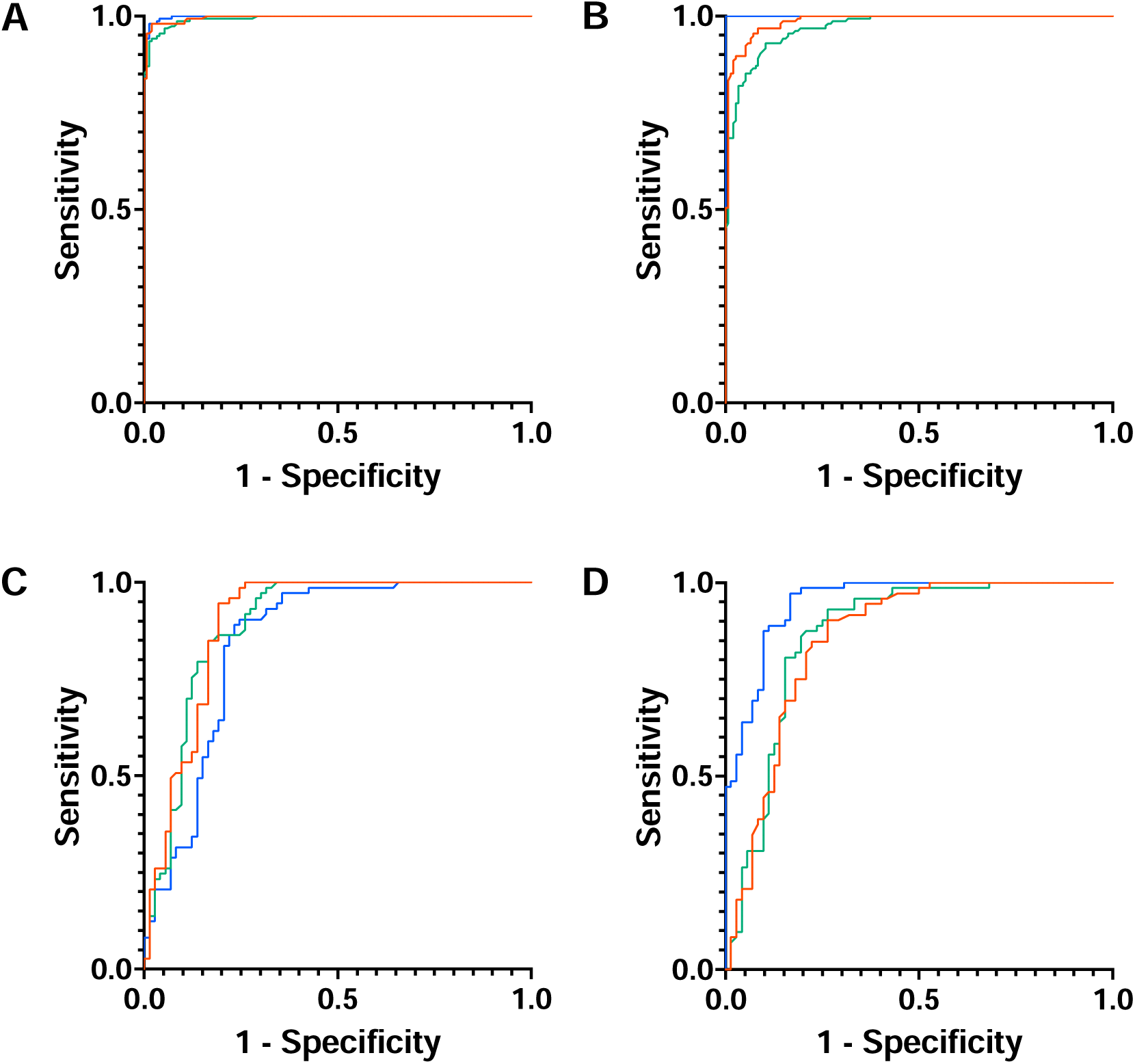
ROC analysis for the discriminative ability of pO_2_ for the identification of adrenal veins. The ability of partial pressure of oxygen (pO_2_) to distinguish right adrenal vein from hepatic vein, or left adrenal vein from inferior phrenic vein, was assessed in panel A-B and C-D, respectively. Panel A (N=154) and C (N=73) illustrate the discriminative ability of pO_2_ at baseline, while panel B (N=155) and D (N=72) show this ability under cosyntropin stimulation, respectively. Blue, red and green lines indicate selectivity index, actual pO_2_ and Δ pO_2_, respectively. Δ pO_2_ was calculated by subtracting pO_2_ levels at external iliac vein from those at a target vein.

**Table 1.**
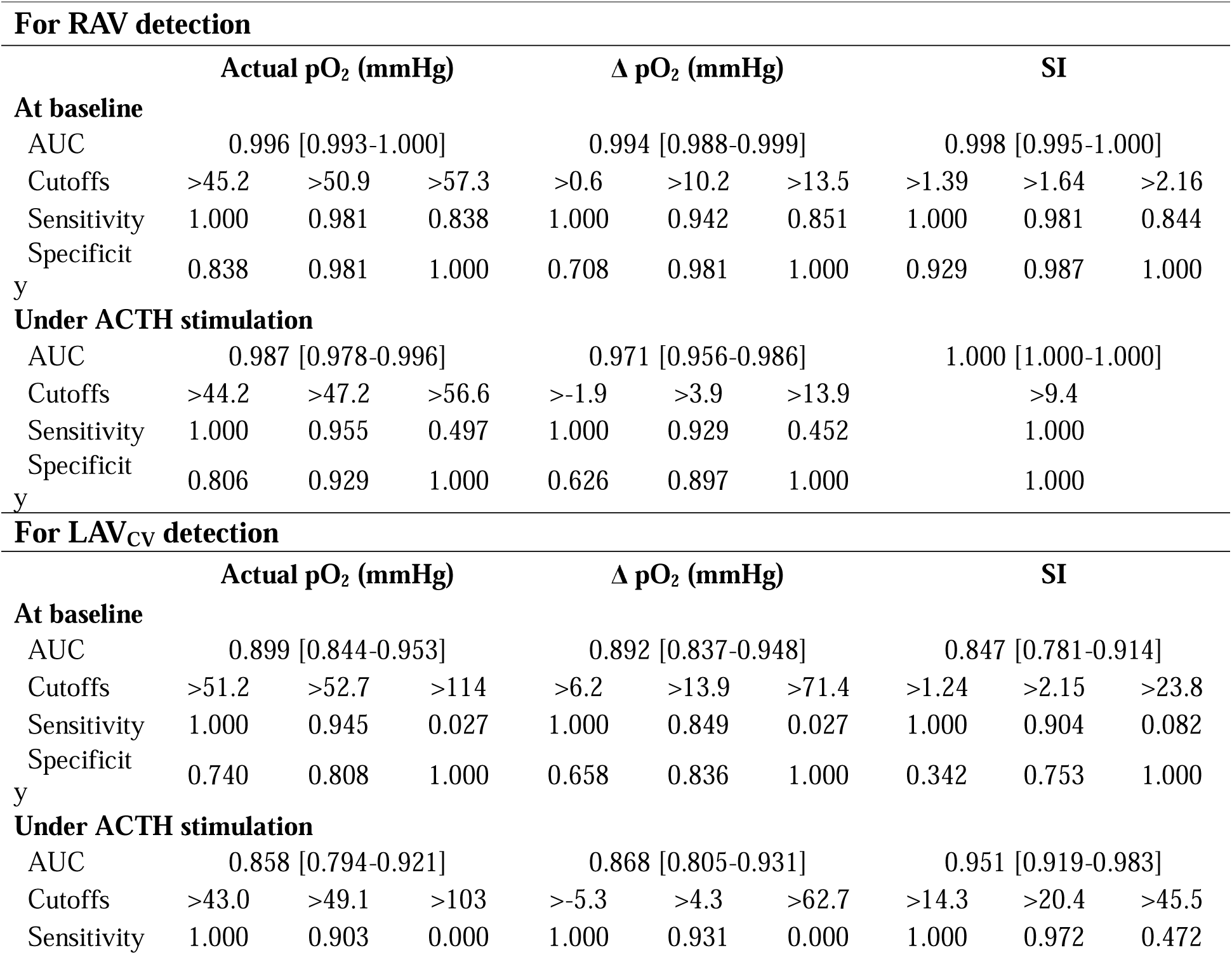
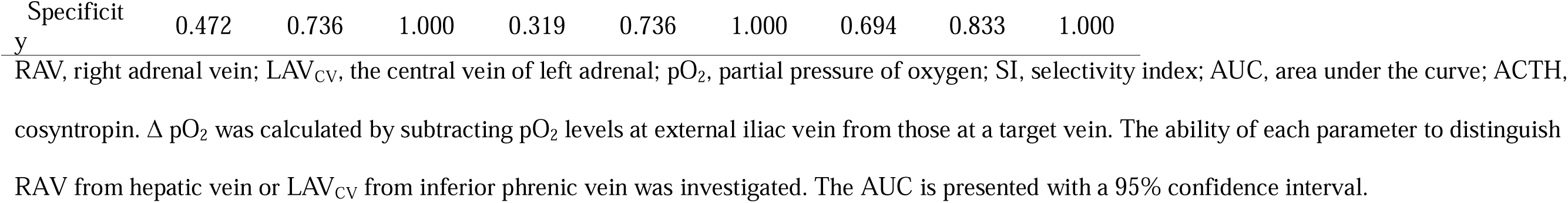
The discriminative ability of partial pressure of oxygen for adrenal veins.

Similarly, actual pO_2_ and ΔpO_2_ were useful indicators for LAV_CV_ cannulation (**Figure 5C and D, and Table 1**). In baseline gas analysis, optimal cutoff values of SI (>2.15), actual pO_2_ (>52.7 mmHg) and ΔpO_2_ (>13.9 mmHg) showed sensitivities of 0.904, 0.945 and 0.849, and specificities of 0.753, 0.808 and 0.836, respectively. Actual pO_2_ and ΔpO_2_ had larger AUCs than the SI (**Figure 5C**), though the differences were not statistically significant. ACTH stimulation clearly enhanced the discriminative ability of the SI in identifying LAV_CV_, while diminishing the performance of actual pO_2_ and ΔpO_2_ (**Figure 5D**). Throughout the investigation of blood gas analysis for AVS, the pCO_2_ and ΔpCO_2_ levels have less discriminative power in distinguishing the adrenal veins from the non-adrenal veins, despite showing significant differences in pCO_2_ between those veins (**Figure S4**).

#### 2. Representative cases illustrating the potential of pO_2_ as a guide in AVS

##### Case 1: Discrimination of RAV from HV

This was the PA case with a common trunk of RAV and HV. Enhanced CT pointed out the possibility of a confluence of RAV and HV prior to AVS (**Figure 6A**). For confirmation of the laterality of PA, we performed AVS in which venous angiography detected a candidate of RAV shown as **Figure 6B**. Blood samples for the assessment of pO_2_ as well as steroids were collected by sequentially inserting a catheter into the superior and inferior branches of the vein to verify whether those veins were draining blood from the right adrenal gland (**Figure 6C and D**). The pO_2_ level was higher in the inferior branch (69.9 mmHg) than in the superior branch (45.1 mmHg) comparable to the trend of SI (**Table 2**). Under ACTH stimulation, the SI significantly elevated in the inferior branch (from 1.76 to 28.8), but not in the superior branch (from 1.09 to 1.34). Taken together with the findings of preoperative CT, we identified the inferior and superior branches as RAV and HV, respectively. The pO_2_ levels maintained their dominance in RAV over HV even after ACTH injection.

**Figure 6.**
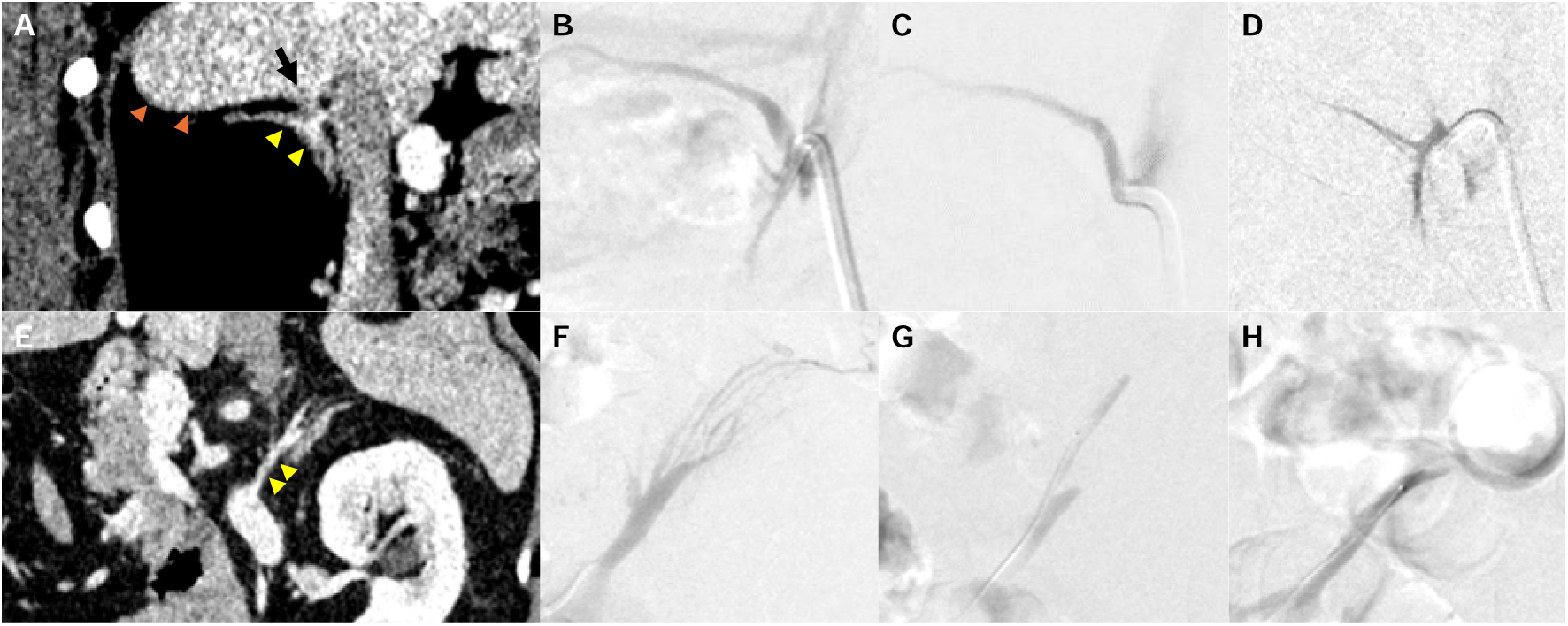
Adrenal imaging and venous angiography of representative cases. Panel A-D and E-H show representative images of computed tomography and venous angiography from two men diagnosed with primary aldosteronism (Case 1 and 2), respectively. In Case 1, preoperative computed tomography revealed a confluence of the right adrenal vein and hepatic vein (A: Yellow and orange arrowheads indicate the right adrenal and the liver, respectively. A black arrow marks the accessory hepatic vein which flows into the right adrenal vein). Adrenal venous sampling confirmed this confluence (B). The right adrenal and hepatic veins were separately selected by a catheter (C and D, respectively) and the venous partial pressure of oxygen levels at baseline were measured as 69.9 and 45.1 mmHg, respectively. In Case 2, enhanced computed tomography visualized the left adrenal vein (E: yellow arrowheads). Adrenal venous sampling identified the left adrenal vein with three main branches (F). After exclusion of the proximal one as inferior phrenic vein, the middle and distal branches were individually cannulated for blood sampling (G and H, respectively), confirming the distal one as the left adrenal central vein based on the selectivity index. The blood partial pressure of oxygen level at baseline was measured as 37.1 mmHg in the middle vein and 57.7 mmHg in the distal vein.

**Table 2.** Venous parameters of representative cases illustrating pO_2_-guided adrenal venous sampling.

| Case 1: Discrimination of RAV from HV |  |  |  |  |  |  |  |  |
| --- | --- | --- | --- | --- | --- | --- | --- | --- |
|  | EIV<br>(reference) |  | HV<br>(shown as Figure 6C) |  | RAV<br>(shown as Figure 6D) |  |  |  |
|  | Baseline | After ACTH | Baseline | After ACTH | Baseline | After ACTH |  |  |
| Cortisol (µg/dL) | 9.6 | 20.9 | 10.5 | 28 | 16.9 | 602 |  |  |
| SI | N/A | N/A | 1.09 | 1.34 | 1.76 | 28.8 |  |  |
| pO<sub>2</sub> (mmHg) | 49.1 | 42.1 | 45.1 | 42.1 | 69.9 | 54.1 |  |  |
| Case 2: Identification of LAV<sub>CV</sub> among a few candidate |  |  |  |  |  |  |  |  |
|  | EIV<br>(reference) |  | LAV<sub>IPV</sub> |  | The middle vein<br>(shown as Figure 6G) |  | The distal vein<br>(shown as Figure 6H) |  |
|  | Baseline | After ACTH | Baseline | After ACTH | Baseline | After ACTH | Baseline | After ACTH |
| Cortisol (µg/dL) | 7.6 | 14.1 | 10.8 | 135 | 8.9 | 32 | 24.5 | 591 |
| SI | N/A | N/A | 1.42 | 9.57 | 1.17 | 2.27 | 3.22 | 41.9 |
| pO<sub>2</sub> (mmHg) | 40.2 | 43 | 44.7 | 47.3 | 37.1 | 35.7 | 57.7 | 53.6 |

RAV, right adrenal vein; HV, hepatic vein; EIV, external iliac vein; ACTH, cosyntropin; SI, selectivity index; pO<sub>2</sub>, partial pressure of oxygen; LAV<sub>CV</sub>, the
central vein of left adrenal; LAV<sub>IPV</sub>, left adrenal vein after the confluence of inferior phrenic vein; IPV, inferior phrenic vein; N/A, not assessed.

##### Case 2: Identification of LAV among several candidate veins

The second case was a PA patient whose adrenal imaging detected a few candidates of LAV_CV_. Preoperative CT with contrast media clearly indicated LAV, with three main branches suspected to drain blood from the left adrenal gland (**Figure 6E**). We considered the proximal one coursing upward as IPV but couldn’t identify which of the two other veins was the LAV_CV_ preoperatively. During AVS, we obtained blood samples from the two veins for the measurement of cortisol and pO_2_ (**Figure 6F**). The SI and pO_2_ levels were summarized in **Table 2**. In the middle branch (**Figure 6G**), the SI and pO_2_ level were 1.17 and 37.1 mmHg, respectively, while those parameters were higher in the distal branch (3.22 and 57.7 mmHg, respectively: **Figure 6H**). After ACTH initiation, the SI increased to 2.27 and 41.9 in the middle and distal ones, respectively, resulting in confirmation of the distal vein as LAV_CV_.

## Discussion

In our present study, we firstly demonstrated the difference of pO_2_ status among the adrenal veins and their surrounding veins, which could be also clinically applicable for identification of adrenal veins in AVS. Of note, pO_2_ status in adrenal veins was significantly negatively associated with steroid production from the adrenal tissue and body mass index. Venous gas analysis in both before and after ACTH stimulation indicated adrenal oxygen consumption for steroid hormone synthesis.

Oxygen is one of the most critical elements for cellular activities. A sufficient supply of oxygen enables living cells to produce adenosine triphosphate, the primary energy source of the cells, and water through cellular respiration occurring in the mitochondria^29^. In the adrenal gland, this aerobic reaction also supports the ACTH-dependent steroidogenesis by enhancing the expression of steroidogenic enzymes using adenosine triphosphate^30^. Results of our present study also demonstrated that pO_2_ levels in the adrenal veins were significantly higher than those in the other veins examined. These findings also suggest that adrenal consumption of oxygen is relatively small, compared to the organs such as the liver, stomach and lower limb. However, the differences in oxygen consumption between the adrenal glands and other organs have been rarely studied in both humans and animals. One exception is an animal study in which adrenal oxygen consumption was slightly lower than that of the liver in rabbits (2.53 ± 0.25 vs. 3.05 ± 0.77 μl/hr/mg dry weight)^31^. From another perspective, the adrenal glands are known to be highly vascularized. In humans, arterial blood supply to bilateral adrenal glands typically originates from the abdominal aorta, renal and phrenic arteries ^32^. Animal studies previously demonstrated that the adrenal glands receive up to 0.5% of the total cardiac output^33–35^, while the adrenal glands are also one of the organs receiving the highest blood flow per unit of weight^34^. Therefore, the adrenal glands have an abundant supply of oxygen. In the adrenal cortex, oxygen is also involved in the steroidogenic process, particularly in oxidation reactions. Several steroidogenic enzymes, including aldosterone synthase, utilize oxygen within the mitochondria and the smooth endoplasmic reticulum during the transformation of cholesterol into steroid hormones^36^. In addition, chromaffin cells in the adrenal medulla also require oxygen for adrenaline synthesis during the hydroxylation step^37^. Those hormonal synthesis also generates reactive oxygen species, which are scavenged by antioxidant enzymes such as copper and zinc and manganese superoxide dismutase (SOD)^38^. In those SOD, copper and zinc-SOD played as a scavenger of toxic superoxide radicals generated during steroidogenesis, while manganese-SOD during catecholamine production^38^. Regarding adrenal oxygen consumption, a couple of animal studies reported that slices of dog and pig adrenal tissues exhibited baseline oxygen uptake rates of 84.4 and 86 mm^3^/100 mg weight/hr at baseline, respectively^39, 40^. Both studies also demonstrated an increased oxygen uptake by 20-40 mm^3^/100 mg weight/hr following administration of ACTH^39, 40^. In addition, oxygen availability has been identified as a regulatory factor in steroidogenesis^41–44^. An *in vitro* study using bovine adrenocortical cells demonstrated that low pO_2_ conditions (80-90 torr) significantly decreased aldosterone production at baseline (-31%) and in response to angiotensin II (-45 to -55%), cAMP (-48 to - 54%) and ACTH (-13 to -33%), compared with normal pO_2_ conditions (140-150 torr)^41^. Similarly, oxygen-dependent regulation of steroidogenesis has also been observed in human tissue experiments, particularly under stimulated conditions^44^. In accordance with those reports, we observed a set of decreased pO_2_ and increased pCO_2_ levels in the adrenal veins after cosyntropin administration. In addition, pO_2_ changes before and after ACTH stimulation were significantly negatively correlated with production of both aldosterone and cortisol in the adrenal veins. Results of our present study are, therefore, interpreted as oxygen consumption during ACTH-dependent steroid synthesis in the adrenal glands. Those findings also suggested that the rich vascularization of the adrenal glands provides sufficient oxygen to support prompt steroid hormone synthesis in response to physiological demand. Consequently, the baseline pO_2_ levels in the adrenal veins were considered higher than in other veins due to the relatively abundant oxygen supply. This characteristic was utilized in the current study to assess successful cannulation to the adrenal veins during AVS. Regarding the observed negative impact of body mass index on adrenal venous pO_2_ levels, several clinical studies reported that obesity was associated with lower oxygen saturation, and lower maximal oxygen uptake during exercise^45–48^. The mechanisms underlying this association has been unclear, but the reduction of expiratory residual volume under obesity is considered as one of the contributing factors^49^. The possible association between adrenal oxygen consumption and the body size has remained unknown at this juncture and therefore, further studies are warranted to expand our understanding about the mechanisms of oxygen consumption in adrenal cortical cells and the influential factors.

Notably, our study firstly indicated that pO_2_ evaluation was a promising tool or pathfinder for identifying adrenal veins during AVS. Nearly 60 years have passed since AVS was first proposed for the subtype determination of PA^50^. However, cannulating the adrenal veins is still clinically challenging. Today, the combination of preprocedual imaging of adrenal veins and development of customized catheter has led to higher AVS success rates than before^15, 28^. Nevertheless, intraprocedural judging whether the cannulated vessel is the target adrenal vein based on the angiography is sometimes difficult, particularly for operators with limited experience^18^. Successful cannulation to the adrenal veins is worldwide defined by achieving a higher SI with specific cutoff values that depend on sampling conditions and regions^17, 20, 21, 28^. Recently, a new assay of cortisol with a shortened measurement time has been developed for the quick assessment of adrenal venous cannulation^18^. This immunochromatographic assay measures plasma cortisol concentrations quantitatively within 6 minutes, increasing successful AVS rates from 54% to 93% judged by the SI in a multi-center study^18^. With the findings from our present study, we propose another and an easy-to-use approach to the judgement of adrenal venous selection using pO_2_ levels in the cannulated vein. Blood gas analyzers are widely available in hospitals, including inpatient unit, catheter laboratory and emergency room. Blood gas analysis needs only a small blood sample and can be completed within one minute. Our findings demonstrated that pO_2_ levels are significantly higher in RAV and LAV_CV_ than in their counterpart veins, HV and IPV, respectively, particularly under the baseline situation. The identical ability of pO_2_ levels for the adrenal veins was compatible to the SI. Thus, AVS operators can easily and repeatedly use the technique to confirm whether the catheter is properly inserted into the adrenal veins until successful cannulation is achieved.

The current study proposes pO_2_-guided AVS and provides a novel insight into adrenal oxygen consumption, while there are a few limitations to be considered for interpreting our findings. First, our AVS protocol includes oxygen administration to settle the patients’ breathing for the efficient performance of AVS. This medical procedure might alter the pO_2_ levels in the examined veins and the cutoff values for the adrenal veins. Second, the cases where we assessed the pO_2_ levels in IPV were limited due to the low clinical necessity, which influence the ROC curve for differentiating LAV_CV_ from IPV. Finally, this is a single-center study, but not a multicenter study. A future study collaborating with multiple international centers is expected to clearly understand the utility of pO_2_ assessment for PA subtyping.

In conclusion, the first awareness of different blood colors between the adrenal and non-adrenal veins brought us to the current study, in which we successfully demonstrated the different distributions of pO_2_ levels in the adrenal and their neighboring veins. Blood gas analysis is a faster, easier and less-expensive tool to identify the adrenal veins during AVS. From bedside to bench, we also observed oxygen consumption in the adrenal glands, referred to as “adrenal breathing”. Our findings should advance both clinical practice and research in the field of adrenal endocrinology.

## Acknowledgments

We sincerely thank Shinichiro Hosaka and Satoko Kurosawa for their clinical management of PA patients. We also appreciate the technical assistance provided by Yasuko Tsukada and Kumi Kikuchi. Additionally, we also extend our heartfelt gratitude to Akane Sugawara and Hiroko Kato for their secretarial assistance.

## Sources of Funding

This work was supported by JSPS KAKENHI Grant Number 23K15091 and YOKOYAMA Foundation for Clinical Pharmacology (YRY-2311).

## Disclosures

KO has received JSPS KAKENHI Grant (23K15091). YT has received JSPS KAKENHI Grant (25K19407) and project support from YOKOYAMA Foundation for Clinical Pharmacology (YRY-2311). YT and YO have received project support from H.U. Group Research Institute and Fujirebio Inc. HK has received research funding from Sumitomo Pharma Co., Ltd., Kyowa Kirin Co., Ltd., Abbott Japan LLC, Nippon Boehringer Ingelheim Co., Ltd. and Murata Manufacturing Co., Ltd.. FS received grant support from the Ministry of Health, Labor and Welfare, Japan (No. H29-Nanji-Ippan-046) and research funding from Nippon Boehringer Ingelheim Co., Ltd.. All other authors declare no conflict of interest.

## Data availability

Data presented in this study are available from the corresponding authors upon request.

## Supplemental Material

Tables S1–S5 Figure S1-S4

## Non-standard Abbreviations and Acronyms

ACTH: adrenocorticotropic hormone
AVS: adrenal venous sampling
BP: blood pressure
CT: computed tomography
CVD: cardiovascular disease
EH: essential hypertension
EIV: external iliac vein
HV: hepatic vein
IVC: inferior vena cava
IPV: inferior phrenic vein
LAV: left adrenal vein
MRA: mineralocorticoid receptor antagonist
OR: odds ratio
PA: primary aldosteronism
PAC: plasma aldosterone concentration
PRA: plasma renin activity
PRC: plasma renin concentration
pCO_2_: partial pressure of carbon dioxide
pO_2_: partial pressure of oxygen
RAV: right adrenal vein
ROC: receiver operating curve
SI: selectivity index
SOD: superoxide dismutase

## References

1. Turcu AF, Yang J and Vaidya A. Primary aldosteronism - a multidimensional syndrome. Nat Rev Endocrinol. 2022;18:665–682.

2. Monticone S, D’Ascenzo F, Moretti C, Williams TA, Veglio F, Gaita F and Mulatero P. Cardiovascular events and target organ damage in primary aldosteronism compared with essential hypertension: a systematic review and meta-analysis. The lancet Diabetes & endocrinology. 2018;6:41–50.

3. Monticone S, Sconfienza E, D’Ascenzo F, Buffolo F, Satoh F, Sechi LA, Veglio F and Mulatero P. Renal damage in primary aldosteronism: a systematic review and meta-analysis. J Hypertens. 2020;38:3–12.

4. Wu X, Yu J and Tian H. Cardiovascular risk in primary aldosteronism: A systematic review and meta-analysis. Medicine. 2019;98:e15985.

5. Meng Z, Dai Z, Huang K, Xu C, Zhang YG, Zheng H and Liu TZ. Long-Term Mortality for Patients of Primary Aldosteronism Compared With Essential Hypertension: A Systematic Review and Meta-Analysis. Front Endocrinol (Lausanne). 2020;11:121.

6. Reincke M, Bancos I, Mulatero P, Scholl UI, Stowasser M and Williams TA. Diagnosis and treatment of primary aldosteronism. The lancet Diabetes & endocrinology. 2021;9:876–892.

7. Marzano L and Ronco C. Clinical and biochemical outcomes after adrenalectomy for primary aldosteronism in tertiary and quaternary referral centers: data from SOPRANO study. Hypertension research : official journal of the Japanese Society of Hypertension. 2024;47:721–734.

8. Hundemer GL, Curhan GC, Yozamp N, Wang M and Vaidya A. Incidence of Atrial Fibrillation and Mineralocorticoid Receptor Activity in Patients With Medically and Surgically Treated Primary Aldosteronism. JAMA Cardiol. 2018;3:768–774.

9. Hundemer GL, Curhan GC, Yozamp N, Wang M and Vaidya A. Cardiometabolic outcomes and mortality in medically treated primary aldosteronism: a retrospective cohort study. The lancet Diabetes & endocrinology. 2018;6:51–59.

10. Chen SY, Chen JY, Huang WC, Puar THK, Chin Kek P, Chueh JS, Lin YH, Wu VC and Study Group T. Cardiovascular outcomes and all-cause mortality in primary aldosteronism after adrenalectomy or mineralocorticoid receptor antagonist treatment: a meta-analysis. Eur J Endocrinol. 2022;187:S47–s58.

11. Tsai CH, Chen YL, Pan CT, Lin YT, Lee PC, Chiu YW, Liao CW, Chen ZW, Chang CC, Chang YY, Hung CS and Lin YH. New-Onset Atrial Fibrillation in Patients With Primary Aldosteronism Receiving Different Treatment Strategies: Systematic Review and Pooled Analysis of Three Studies. Front Endocrinol (Lausanne). 2021;12:646933.

12. Rossi GP, Rossitto G, Amar L, Azizi M, Riester A, Reincke M, Degenhart C, Widimsky J, Jr., Naruse M, Deinum J, Schultze Kool L, Kocjan T, Negro A, Rossi E, Kline G, Tanabe A, Satoh F, Christian Rump L, Vonend O, Willenberg HS, Fuller PJ, Yang J, Chee NYN, Magill SB, Shafigullina Z, Quinkler M, Oliveras A, Dun Wu K, Wu VC, Kratka Z, Barbiero G, Battistel M, Chang CC, Vanderriele PE and Pessina AC. Clinical Outcomes of 1625 Patients With Primary Aldosteronism Subtyped With Adrenal Vein Sampling. Hypertension (Dallas, Tex : 1979). 2019;74:800–808.

13. Chayovan T, Limumpornpetch P and Hongsakul K. Success rate of adrenal venous sampling and predictors for success: a retrospective study. Pol J Radiol. 2019;84:e136–e141.

14. Muhamad Pauzi KN, Zakaria R, Leong YY, Nik Fuad NF, Nik Ismail NA and Sukor N. Success Rate of Adrenal Venous Sampling and its Determining Factors: Experience of a Single Center in Malaysia. Ann Vasc Surg. 2024;98:258–267.

15. Omura K, Ota H, Takahashi Y, Matsuura T, Seiji K, Arai Y, Morimoto R, Satoh F and Takase K. Anatomical Variations of the Right Adrenal Vein: Concordance Between Multidetector Computed Tomography and Catheter Venography. *Hypertension (Dallas*, Tex : *1979*). 2017;69:428–434.

16. Araki T, Imaizumi A, Okada H, Sasaki Y and Onishi H. Anatomy of Left Inferior Phrenic Vein in Patients Without Portal Hypertension. AJR Am J Roentgenol. 2021;217:411–417.

17. Rossitto G, Amar L, Azizi M, Riester A, Reincke M, Degenhart C, Widimsky J, Naruse M, Deinum J, Schultzekool L, Kocjan T, Negro A, Rossi E, Kline G, Tanabe A, Satoh F, Rump LC, Vonend O, Willenberg HS, Fuller P, Yang J, Nian Chee NY, Magill SB, Shafigullina Z, Quinkler M, Oliveras A, Chang CC, Wu VC, Somloova Z, Maiolino G, Barbiero G, Battistel M, Lenzini L, Quaia E, Pessina AC and Rossi GP. Subtyping of Primary Aldosteronism in the AVIS-2 Study: Assessment of Selectivity and Lateralization. The Journal of clinical endocrinology and metabolism. 2020;105.

18. Yoneda T, Karashima S, Kometani M, Usukura M, Demura M, Sanada J, Minami T, Koda W, Gabata T, Matsui O, Idegami K, Takamura Y, Tamiya E, Oe M, Nakai M, Mori S, Terayama N, Matsuda Y, Kamemura K, Fujii S, Seta T, Sawamura T, Okuda R, Takeda Y, Hayashi K, Yamagishi M and Takeda Y. Impact of New Quick Gold Nanoparticle-Based Cortisol Assay During Adrenal Vein Sampling for Primary Aldosteronism. The Journal of clinical endocrinology and metabolism. 2016;101:2554–61.

19. Takahashi Y, Ota H, Omura K, Dendo Y, Otani K, Matsuura T, Kitami M, Seiji K, Tezuka Y, Nezu M, Ono Y, Morimoto R, Satoh F and Takase K. Image quality and radiation dose of low-tube-voltage CT with reduced contrast media for right adrenal vein imaging. Eur J Radiol. 2018;98:150–157.

20. Naruse M, Katabami T, Shibata H, Sone M, Takahashi K, Tanabe A, Izawa S, Ichijo T, Otsuki M, Omura M, Ogawa Y, Oki Y, Kurihara I, Kobayashi H, Sakamoto R, Satoh F, Takeda Y, Tanaka T, Tamura K, Tsuiki M, Hashimoto S, Hasegawa T, Yoshimoto T, Yoneda T, Yamamoto K, Rakugi H, Wada N, Saiki A, Ohno Y and Haze T. Japan Endocrine Society clinical practice guideline for the diagnosis and management of primary aldosteronism 2021. Endocrine journal. 2022;69:327–359.

21. Funder JW, Carey RM, Mantero F, Murad MH, Reincke M, Shibata H, Stowasser M and Young WF, Jr. The Management of Primary Aldosteronism: Case Detection, Diagnosis, and Treatment: An Endocrine Society Clinical Practice Guideline. The Journal of clinical endocrinology and metabolism. 2016;101:1889–916.

22. Ono Y, Tezuka Y, Omata K, Morimoto R, Yamazaki Y, Oguro S, Takase K, Ito A, Yoshimi T, Kojima S, Ito S, Sasano H, Suzuki T, Tanaka T, Katagiri H and Satoh F. Screening Cutoff Values for the Detection of Aldosterone-Producing Adenoma by LC-MS/MS and a Novel Noncompetitive CLEIA. Journal of the Endocrine Society. 2024;8:bvae080.

23. Tezuka Y, Omata K, Ono Y, Kambara K, Kamada H, Oguro S, Yamazaki Y, Gomez-Sanchez CE, Ito A, Sasano H, Takase K, Tanaka T, Katagiri H and Satoh F. Investigating the cut-off values of captopril challenge test for primary aldosteronism using the novel chemiluminescent enzyme immunoassay method: a retrospective cohort study. Hypertension research : official journal of the Japanese Society of Hypertension. 2024;47:1362–1371.

24. Reincke M. Subclinical Cushing’s syndrome. Endocrinology and metabolism clinics of North America. 2000;29:43–56.

25. Satani N, Ota H, Seiji K, Morimoto R, Kudo M, Iwakura Y, Ono Y, Nezu M, Omata K, Ito S, Satoh F and Takase K. Intra-adrenal Aldosterone Secretion: Segmental Adrenal Venous Sampling for Localization. Radiology. 2016;278:265–74.

26. Makita K, Nishimoto K, Kiriyama-Kitamoto K, Karashima S, Seki T, Yasuda M, Matsui S, Omura M and Nishikawa T. A Novel Method: Super-selective Adrenal Venous Sampling. J Vis Exp. 2017.

27. Tannai H, Makita K, Koike Y, Nakai K, Tsurutani Y, Okudela K, Saito J, Matsui S, Kakuta Y and Nishikawa T. Usefulness and accuracy of segmental adrenal venous sampling on localisation and functional diagnosis of various adrenal lesions in primary aldosteronism. Clin Radiol. 2022;77:e652–e659.

28. Satoh F, Morimoto R, Seiji K, Satani N, Ota H, Iwakura Y, Ono Y, Kudo M, Nezu M, Omata K, Tezuka Y, Kawasaki Y, Ishidoya S, Arai Y, Takase K, Nakamura Y, McNamara K, Sasano H and Ito S. Is there a role for segmental adrenal venous sampling and adrenal sparing surgery in patients with primary aldosteronism? Eur J Endocrinol. 2015;173:465–77.

29. Babcock GT. How oxygen is activated and reduced in respiration. Proc Natl Acad Sci U S A. 1999;96:12971–3.

30. Miller WL. Steroid hormone synthesis in mitochondria. Molecular and cellular endocrinology. 2013;379:62–73.

31. Rosenkrantz H. Studies in vitamin E deficiency. I. The oxygen consumption of various tissues from the rabbit. J Biol Chem. 1955;214:789–97.

32. Abdellatif AB, Fernandes-Rosa FL, Boulkroun S and Zennaro MC. Vascular and hormonal interactions in the adrenal gland. Front Endocrinol (Lausanne). 2022;13:995228.

33. Lundeen G, Manohar M and Parks C. Systemic distribution of blood flow in swine while awake and during 1.0 and 1.5 MAC isoflurane anesthesia with or without 50% nitrous oxide. Anesth Analg. 1983;62:499–512.

34. Johnston BM and Owen DA. Tissue blood flow and distribution of cardiac output in cats: changes caused by intravenous infusions of histamine and histamine receptor agonists. Br J Pharmacol. 1977;60:173–80.

35. Gomez-Sanchez CE. Regulation of adrenal arterial tone by adrenocorticotropin: the plot thickens. Endocrinology. 2007;148:3566–8.

36. Nishimoto K, Ogishima T, Sugiura Y, Suematsu M and Mukai K. Pathology and gene mutations of aldosterone-producing lesions. Endocrine journal. 2023;70:1113–1122.

37. Berends AMA, Eisenhofer G, Fishbein L, Horst-Schrivers A, Kema IP, Links TP, Lenders JWM and Kerstens MN. Intricacies of the Molecular Machinery of Catecholamine Biosynthesis and Secretion by Chromaffin Cells of the Normal Adrenal Medulla and in Pheochromocytoma and Paraganglioma. Cancers (Basel). 2019;11.

38. Sasano H, Mizorogi A, Sato M, Nakazumi H and Suzuki T. Superoxide Dismutase in Human Adrenal and its Disorders: A Correlation with Development and Neoplastic Changes. Endocr Pathol. 1999;10:325–333.

39. McKee RW and Walker JK. Oxygen consumption of adrenal slices from normal and scorbutic guinea pigs and the influence of added ACTH. *Science (New York*, NY). 1953;118:133–5.

40. Nichols J, Davis C and Green HD. Effect of hypophysectomy, DDD treatment, and surgical trauma on the oxygen consumption of the various zones of the adrenal cortex. Endocrinology. 1953;53:541–8.

41. Raff H, Ball DL and Goodfriend TL. Low oxygen selectively inhibits aldosterone secretion from bovine adrenocortical cells in vitro. Am J Physiol. 1989;256:E640–4.

42. Chabre O, Defaye G and Chambaz EM. Oxygen availability as a regulatory factor of androgen synthesis by adrenocortical cells. Endocrinology. 1993;132:255–60.

43. Stevens VL, Aw TY, Jones DP and Lambeth JD. Oxygen dependence of adrenal cortex cholesterol side chain cleavage. Implications in the rate-limiting steps in steroidogenesis. J Biol Chem. 1984;259:1174–9.

44. Raff H and Bruder ED. Steroidogenesis in human aldosterone-secreting adenomas and adrenal hyperplasias: effects of hypoxia in vitro. Am J Physiol Endocrinol Metab. 2006;290:E199–e203.

45. Ceylan B, Khorshid L, Güneş Ü Y and Zaybak A. Evaluation of oxygen saturation values in different body positions in healthy individuals. J Clin Nurs. 2016;25:1095–100.

46. Jalili M, Nazem F, Sazvar A and Ranjbar K. Prediction of Maximal Oxygen Uptake by Six-Minute Walk Test and Body Mass Index in Healthy Boys. J Pediatr. 2018;200:155–159.

47. Kabon B, Nagele A, Reddy D, Eagon C, Fleshman JW, Sessler DI and Kurz A. Obesity decreases perioperative tissue oxygenation. Anesthesiology. 2004;100:274–80.

48. Vignati C, Mapelli M, Nusca B, Bonomi A, Salvioni E, Mattavelli I, Sciomer S, Faini A, Parati G and Agostoni P. A Breathtaking Lift: Sex and Body Mass Index Differences in Cardiopulmonary Response in a Large Cohort of Unselected Subjects with Acute Exposure to High Altitude. High Alt Med Biol. 2021;22:379–385.

49. Littleton SW and Tulaimat A. The effects of obesity on lung volumes and oxygenation. Respir Med. 2017;124:15–20.

50. Melby JC, Spark RF, Dale SL, Egdahl RH and Kahn PC. Diagnosis and localization of aldosterone-producing adenomas by adrenal-vein catheterization. N Engl J Med. 1967;277:1050–6.

